# Altered cortical thickness development in 22q11.2 deletion syndrome and association with psychotic symptoms

**DOI:** 10.1101/2020.11.03.20221978

**Authors:** Joëlle Bagautdinova, Daniela Zöller, Marie Schaer, Maria Carmela Padula, Valentina Mancini, Maude Schneider, Stephan Eliez

## Abstract

Schizophrenia has been extensively associated with reduced cortical thickness (CT), and its neurodevelopmental origin is increasingly acknowledged. However, the exact timing and extent of alterations occurring in preclinical phases remain unclear. With a high prevalence of psychosis, 22q11.2 deletion syndrome (22q11DS) is a neurogenetic disorder that represents a unique opportunity to examine brain maturation in high-risk individuals. In this study, we quantified trajectories of CT maturation in 22q11DS and examined the association of CT development with the emergence of psychotic symptoms. Longitudinal structural MRI data with 1-6 time points were collected from 324 participants aged 5-35 years (N=148 22q11DS, N=176 controls), resulting in a total of 636 scans (N=334 22q11DS, N=302 controls). Mixed model regression analyses were used to compare CT trajectories between participants with 22q11DS and controls. Further, CT trajectories were compared between participants with 22q11DS who developed (N=61, 146 scans), or remained exempt of (N=47; 98 scans) positive psychotic symptoms during development. Compared to controls, participants with 22q11DS showed widespread increased CT, focal reductions in the posterior cingulate gyrus and superior temporal gyrus (STG), and accelerated cortical thinning during adolescence, mainly in fronto-temporal regions. Within 22q11DS, individuals who developed psychotic symptoms showed exacerbated cortical thinning in the right STG. Together, these findings suggest that genetic predisposition for psychosis is associated with increased CT starting from childhood and altered maturational trajectories of CT during adolescence, affecting predominantly fronto-temporal regions. In addition, accelerated thinning in the STG may represent an early biomarker associated with the emergence of psychotic symptoms.

## Introduction

In recent years, considerable research efforts in schizophrenia have provided extensive support for a neurodevelopmental origin of the illness involving altered grey matter structure. Studies consistently report reduced volume and cortical thickness (CT) in frontal, temporal and cingulate areas (1–6), suggesting alterations in the neuropil of affected individuals. Post-mortem studies revealing decreased synaptic density in brain tissue of patients with schizophrenia provide additional supportive evidence (reviewed in (7)). Importantly, findings suggest that structural alterations appear in a progressive manner, with milder grey matter reductions in individuals at clinical high risk and gradually more aggravated differences in patients with first-episode psychosis and chronic schizophrenia (4,8,9).

While the neurodevelopmental origin of schizophrenia is therefore increasingly validated, gaining insights into the period that precedes the onset of the disorder remains challenging. Prospective studies on ultra-high-risk individuals have emerged to address this question and have shown steeper rates of cortical thinning in individuals who eventually convert to psychosis, affecting fronto-temporal regions (9–11), as well as cingulate, parietal and insular areas (11). However, studies investigating UHR individuals have been limited by their number of follow-up scans (e.g., 1 follow-up) and did not assess neurodevelopment in at-risk individuals before the onset of psychotic symptoms.

With a 30-40% conversion rate to psychosis, 22q11.2 Deletion Syndrome (22q11DS) is considered a genetic model to study the developmental factors related to the emergence of psychosis (12,13). The syndrome is caused by a deletion of 1.5-3Mb on the long arm of chromosome 22. Syndromic individuals are typically identified very early during development and can therefore be followed-up starting from childhood, providing a unique opportunity to gain insight into the neurodevelopmental mechanisms preceding the onset of psychosis. Early neuroimaging studies in 22q11DS reported volumetric reductions in posterior regions, with a relative preservation of frontal volumes (14–18). Cross-sectional studies focusing on more fine-grained, surface-based measures of CT reported widespread increased CT in 22q11DS and focal decreases in the posterior cingulate and superior temporal gyrus (19–21), a finding that was recently confirmed in a large multisite study (22). Some cross-sectional studies furthermore suggested the presence of potentially altered grey matter development, although the extent of developmental alterations is unclear, as some reported faster thinning (23), while others found a lack of thinning (21) or no difference in thinning (22) in 22q11DS compared to controls.

However, given the compelling evidence for complex structural changes during normative brain development (24), cross-sectional studies may not fully capture the extent of alterations in 22q11DS. Few longitudinal studies have provided preliminary evidence into the developmental profile of 22q11DS, but findings remain inconsistent: volumetric studies reported increases (25) or no change (26) of grey matter volume with age in frontal regions, and a decline in occipito-parietal regions (25); CT studies further showed both slower thinning in left parietal regions (27) and accelerated thinning in fronto-temporal regions (23). Finally, the presence of psychotic symptoms has been associated with steeper grey matter decline in frontal (26,28,29), temporal (26), cingulate (29) and parietal regions (25,29). The heterogeneity of findings from longitudinal studies in 22q11DS may be explained by their limited sample size (N=44-135) and follow-up assessments (typically 2-3 scans), preventing a precise delineation of the topology and timing of CT alterations in 22q11DS. A large-scale longitudinal study investigating grey matter development with a fine-grained temporal and topological resolution is thus critical to achieve a more robust and accurate understanding of neurodevelopmental mechanisms that underlie genetic vulnerability for psychosis.

Hence, using the largest-ever longitudinal data set of individuals with 22q11DS and controls comprising 636 scans with up to 6 assessments covering an age span of 5 to 35 years, our first objective was to provide a robust characterization of developmental trajectories of CT in 22q11DS and controls. Next, using a subset of 244 scans from individuals with 22q11DS with up to 5 assessments covering an age span of 6 to 28 years, our second objective was to determine whether the onset of positive psychotic symptoms is characterized by a specific CT maturational profile. Mixed models regression was applied at each cortical vertex to accurately capture developmental trajectories with a high topological precision.

## Method

### Participants

Participants were recruited in the context of an ongoing longitudinal study, through parent associations and word of mouth (23,30). Written informed consent was received from participants and their parents (for subjects <18 years old), under protocols approved by the Cantonal Research Ethics Commission. The presence of a 22q11.2 microdeletion was confirmed using Quantitative Fluorescent Polymerase Chain Reaction (QF-PCR).

For the modeling of CT development in 22q11DS and controls, the sample consisted of 324 participants (N=148 22q11DS (50.7% female); N=176 controls (58.1% female)) aged 5-35 years who contributed 1-6 scans, resulting in a total of 636 scans (N=334 22q11DS; N=302 controls) (Figure 1A). Controls were carefully screened to exclude any neurological or psychiatric disorder. Next, to investigate the association of CT maturation with the emergence of positive psychotic symptoms, we selected a subsample of participants with 22q11DS who underwent the Structured Interview for Psychosis-Risk Syndromes (SIPS) (31). Specifically, participants were classified as having “lifetime attenuated psychotic symptoms” (LA-PS) when they fulfilled the attenuated positive symptoms criterion of the ultra-high-risk status, defined as a score ≥3 at any of the positive symptoms subscales P1-P5 (P1: unusual thought content/delusional ideas, P2: suspiciousness/persecutory ideas, P3: grandiose ideas, P4: perceptual abnormalities/hallucinations, P5: disorganized communication) at any of their assessments. Otherwise, they were classified as having “no psychotic symptoms” (N-PS). The sample selected based on these criteria consisted of 108 participants with 22q11DS (N=47 N-PS, 48.9% female; N=61 LA-PS, 47.5% female) aged 6-28 years who contributed 1-5 scans, resulting in a total of 244 scans (N=98 N-PS; N=146 LA-PS) (Figure 1B). In addition, to assess CT maturation in individuals with 22q11DS who develop overt psychosis, we conducted an exploratory analysis comparing three subgroups: individuals with low psychotic symptoms (N-PS, N=47), individuals with psychotic symptoms who did not develop psychosis (LA-PS no psychosis, N=49), and individuals with psychotic symptoms and a diagnosis of psychosis (LA-PS with psychosis, N=12) (Figure S1). Presence of psychosis was established when participants fulfilled criteria for diagnosis at any of their time points. Details on participant demographics, neurocognitive measures and psychiatric assessment can be found in the Supplementary Method and Table S1.

**Figure 1.**
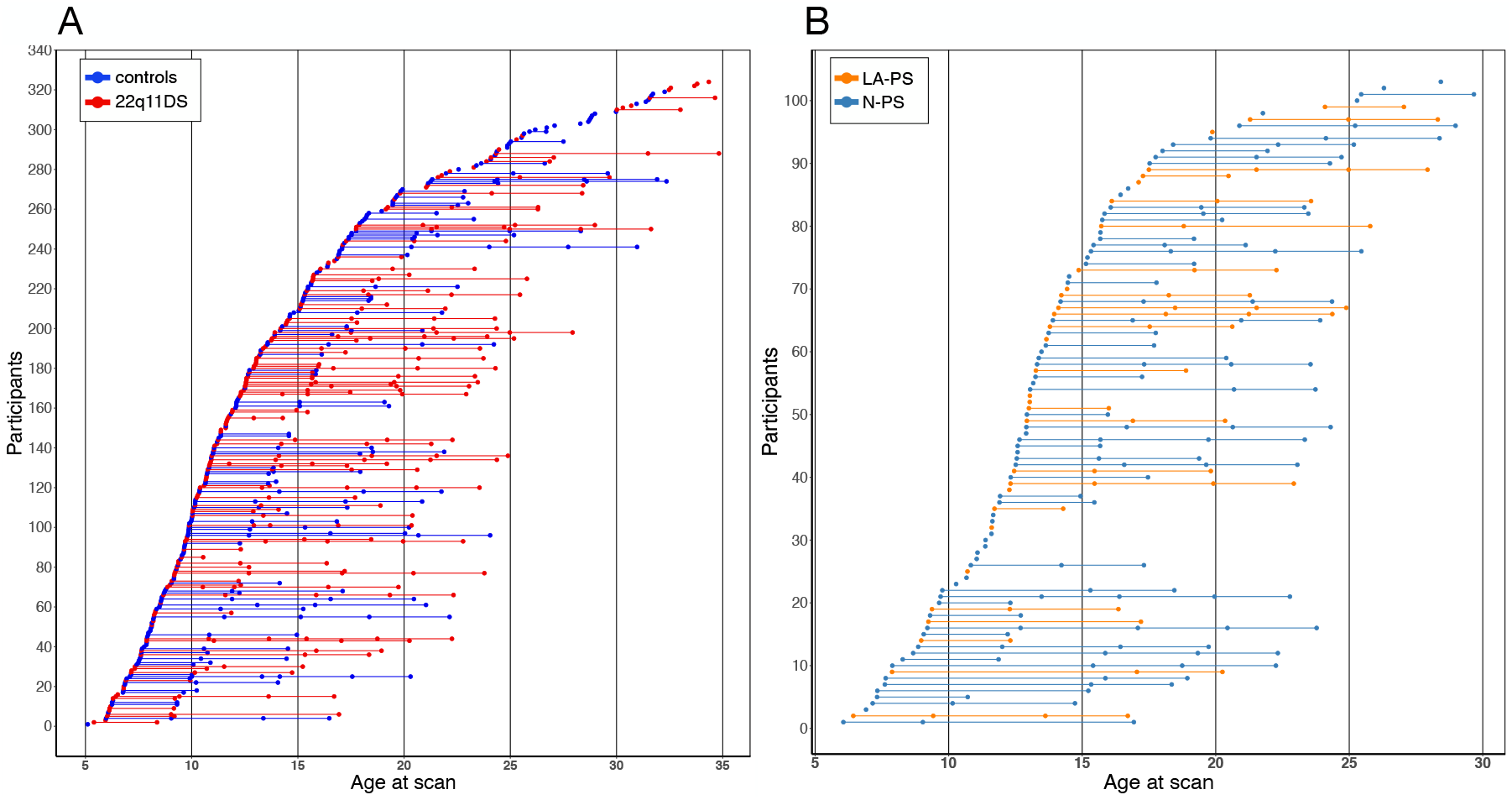
Scan distribution for the analyses comparing A) 22q11DS and controls and B) LA-PS vs N-PS subgroups within 22q11DS. For 22q11DS and control groups, the sample consisted of 324 participants (N=148 22q11DS (73 male, 75 female); N = 176 controls (90 male, 86 female)) aged 5-35 years who contributed between 1-6 scans, resulting in a total of 636 scans (N=334 22q11DS (153 male, 181 female); N = 302 controls (143 male, 159 female). For LA-PS and N-PS groups, the sample consisted of 108 individuals with 22q11DS (N=47 N-PS (24 male, 23 female); N=61 LA-PS (32 male, 29 female)) aged 6-28 years who contributed between 1-5 scans, resulting in a total of 244 scans (N=98 N-PS (48 male, 50 female); N=146 LA-PS (68 male, 78 female).

### Imaging

T1-weighted MRI anatomical brain scans were acquired using three different scanners: a 1.5T Philips Intera scanner (158 scans), a 3T Siemens Trio scanner (300 scans) and a 3T Siemens Prisma scanner (178 scans); acquisition parameters are reported in the Supplementary method. The proportion of scans acquired with each scanner did not differ between 22q11DS and controls (p=0.07) or LA-PS and N-PS groups (p=0.83).

Anatomical segmentation of T1-weighted images was performed using FreeSurfer v5.1 (http://surfer.nmr.mgh.harvard.edu) and cortical thickness (CT) was computed at each vertex. Details of image acquisition and processing are reported in the Supplementary Method.

### Statistical Analyses

Mixed models regression analyses were used to characterize CT maturation over time at each vertex using MATLAB R2017a (MathWorks) (using code published at: https://github.com/danizoeller/myMixedModelsTrajectories) and GNU Parallel (32). This method has been applied in previous studies by our group with a similar longitudinal design (29,30,33– 35), as this approach is suitable for longitudinal datasets such as ours, with variability in the amount of scans per individual and in time between scans. Population parameters (group, age and their interaction) were modeled as fixed effects, and subjects were modeled as random effects. The following equation was used to model CT as a function of age and group at each vertex:

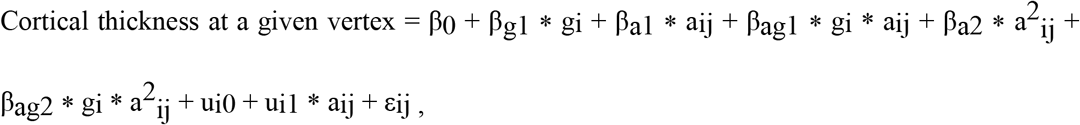

where i = subject index; j = scan index; β_xn_ = fixed effects; g = grouping variable, a = age; u = normally distributed random effect; and ε = normally distributed error term.

Sex and scanner type were included as covariates. Constant (no age-dependence, βa1=βag1=βa2=βag2=0), linear (βa2= βag2=0) and quadratic (full equation above) random-slope models were fitted to the CT data at each vertex for the 22q11DS vs controls comparison; constant and linear models were fitted for the comparison of subgroups within 22q11DS to prevent overfitting in reduced samples. The best model order for a given vertex was determined using the Bayesian Information Criterion (BIC).

Then, the significance of group (intercept) and interaction (shape) effects was assessed using a log-likelihood ratio test comparing the full model described above with reduced models lacking group or age by diagnosis interaction terms, yielding respectively a model without group differences or with similar curve shapes in both groups. For all analyses, cluster-wise correction for multiple comparisons was applied at the significance threshold of p<0.05 using Monte-Carlo permutation (36). Result figures contain CT trajectories of the vertex with the largest effect within significant clusters.

Finally, to capture the timing of CT differences between groups (22q11DS *vs* controls; LA-PS *vs* N-PS), we computed snapshots of 1) the difference in CT between the estimated trajectories of each group, and 2) the difference in the annual rate of cortical thinning of each group at different ages. Corresponding videos showing dynamic CT and thinning differences between groups throughout development are available in the Supplementary Materials.

## Results

### Increased cortical thickness and accelerated thinning in 22q11DS

Vertex-wise mixed models regression of CT development in individuals with 22q11Ds and controls yielded mostly linear (predominantly in frontal and temporal areas) or quadratic (prefrontal and parietal regions) model fits, with some constant model fits (occipital, precentral and medial temporal regions). In both groups, CT trajectories were generally decreasing throughout childhood and adolescence, and either continued decreasing or leveled out at a minimum during adulthood (Figure 2).

**Figure 2.**
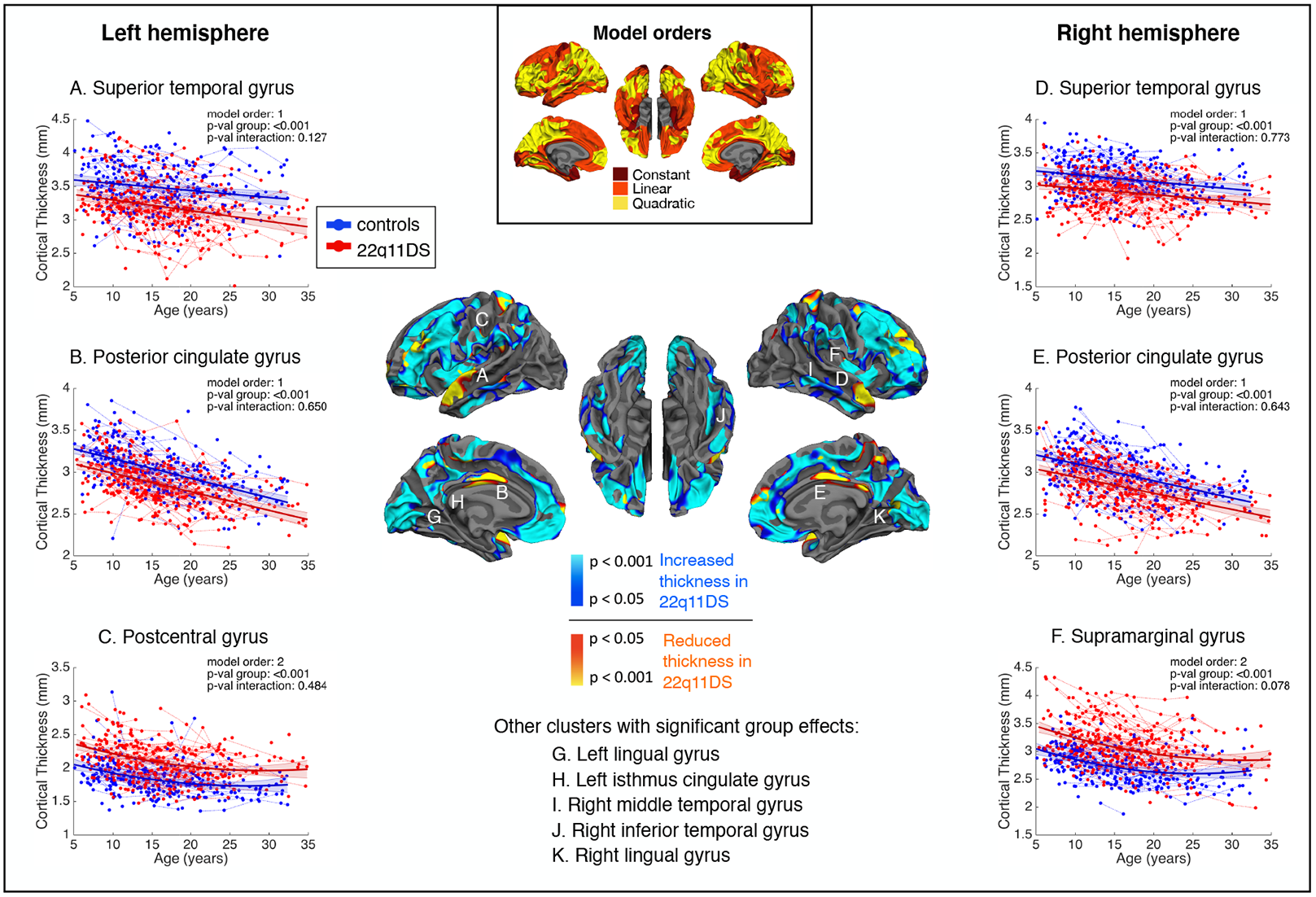
Significant intercept differences in cortical thickness between 22q11DS and controls after cluster-wise correction for multiple comparisons. In the central map, cold colors reflect increased cortical thickness and warm colors indicate reduced cortical thickness in 22q11DS compared to controls. Individuals with 22q11DS show widespread increases in cortical thickness, with focal reductions in the superior temporal gyrus (STG) and in the posterior cingulate cortex (PCC). Representative thinning trajectories are displayed for the bilateral STG and PCC, the left postcentral gyrus and the right supramarginal gyrus (legends A-F). For graphical representations of trajectories from other significant clusters (legends G-K), see Supplementary Figure S3. The middle upper map depicts model orders fitted at each vertex, with dark red indicating constant models, orange corresponding to linear models and yellow indicating quadratic models.

Group differences were widespread and involved higher CT in individuals with 22q11DS through most cortical regions, with cluster peaks located in the bilateral lingual gyri, left postcentral and isthmus cingulate gyri and right supramarginal, middle and inferior temporal gyri. Notable exceptions were observed in the bilateral posterior cingulate cortex (PCC) and superior temporal gyri (STG), where CT was significantly lower in participants with 22q11DS compared to controls.

Moreover, significant group by age interaction effects were evident in several brain regions, reflecting aberrant CT maturation in 22q11DS (Figure 3). In these regions, participants with 22q11DS displayed abnormally higher CT values during childhood, followed by accelerated rates of cortical thinning, such that trajectories either converged towards normative development in adulthood, or continued decreasing. Cluster peaks of altered developmental trajectories were predominantly found in fronto-temporal regions including the left rostral middle, superior frontal, lateral orbitofrontal gyri, right paracentral and bilateral precentral gyri as well left middle temporal, fusiform and parahippocampal regions and the right inferior temporal gyrus. Additional clusters of altered CT trajectories were present in the left superior parietal and the right insula, lateral occipital and lingual regions. Supplementary Table S2 and Figures S2, S3 contain detailed information of each of the clusters showing significant group or interaction effects.

**Figure 3.**
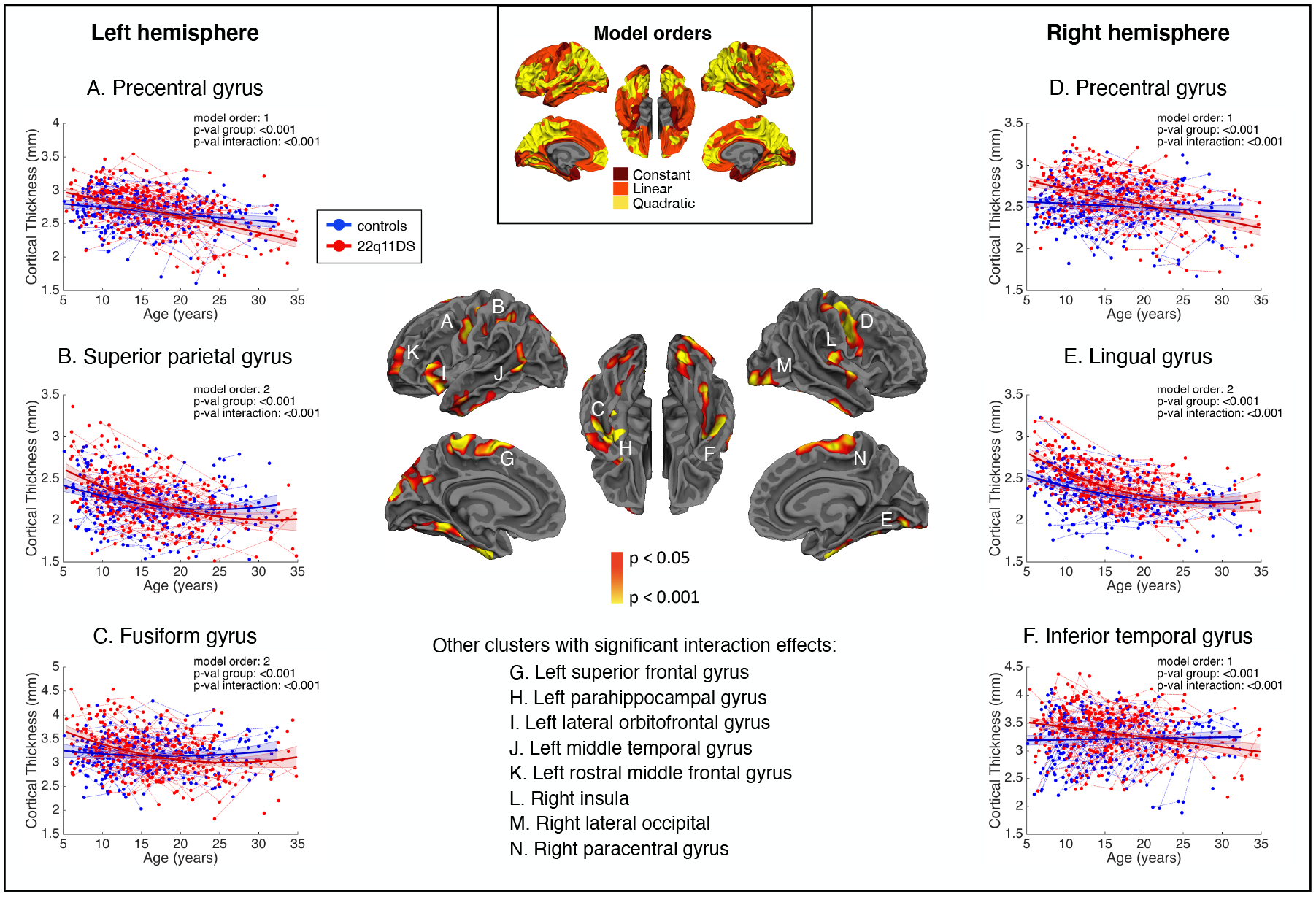
Significant shape differences in cortical thickness between 22q11DS and controls after cluster-wise correction for multiple comparisons. In the central map, warm colors reflect the degree of significance of shape differences found at each vertex. Individuals with 22q11DS show increased cortical thickness during childhood followed by accelerated thinning during adolescence, affecting most prominently fronto-temporal regions. Representative thinning trajectories are displayed for the bilateral precentral gyrus, left superior parietal gyrus, left fusiform gyrus, right lingual gyrus and right inferior temporal gyrus (legends A-F). For graphical representations of trajectories from other significant clusters (legends G-N), see Supplementary Figure S4. The middle upper map depicts model orders fitted at each vertex, with dark red indicating constant models, orange corresponding to linear models and yellow indicating quadratic models.

Snapshots of differences in CT between both groups shown throughout entire age range show left hemisphere regions in children with 22q11DS with a thicker cortex compared to controls (Figure 4A). CT differences progressively lessen throughout adolescence and become less widespread towards adulthood. Differences in annualized thinning rates confirmed that individuals with 22q11DS had faster rates of cortical thinning compared to controls throughout childhood and adolescence (Figure 4B). Interestingly, thinning rates become particularly exacerbated in certain regions when entering adulthood (e.g., left frontal and parietal regions, bilateral insula) reflecting a continued thinning process in individuals with 22q11DS that contrasts with healthy adults, where CT tends to stabilize. For corresponding brain maps of the right hemisphere, see Supplementary Figure S4; Supplementary Videos 1 and 2 show dynamic changes over time of CT and thinning rate differences between 22q11DS and controls.

**Figure 4.**
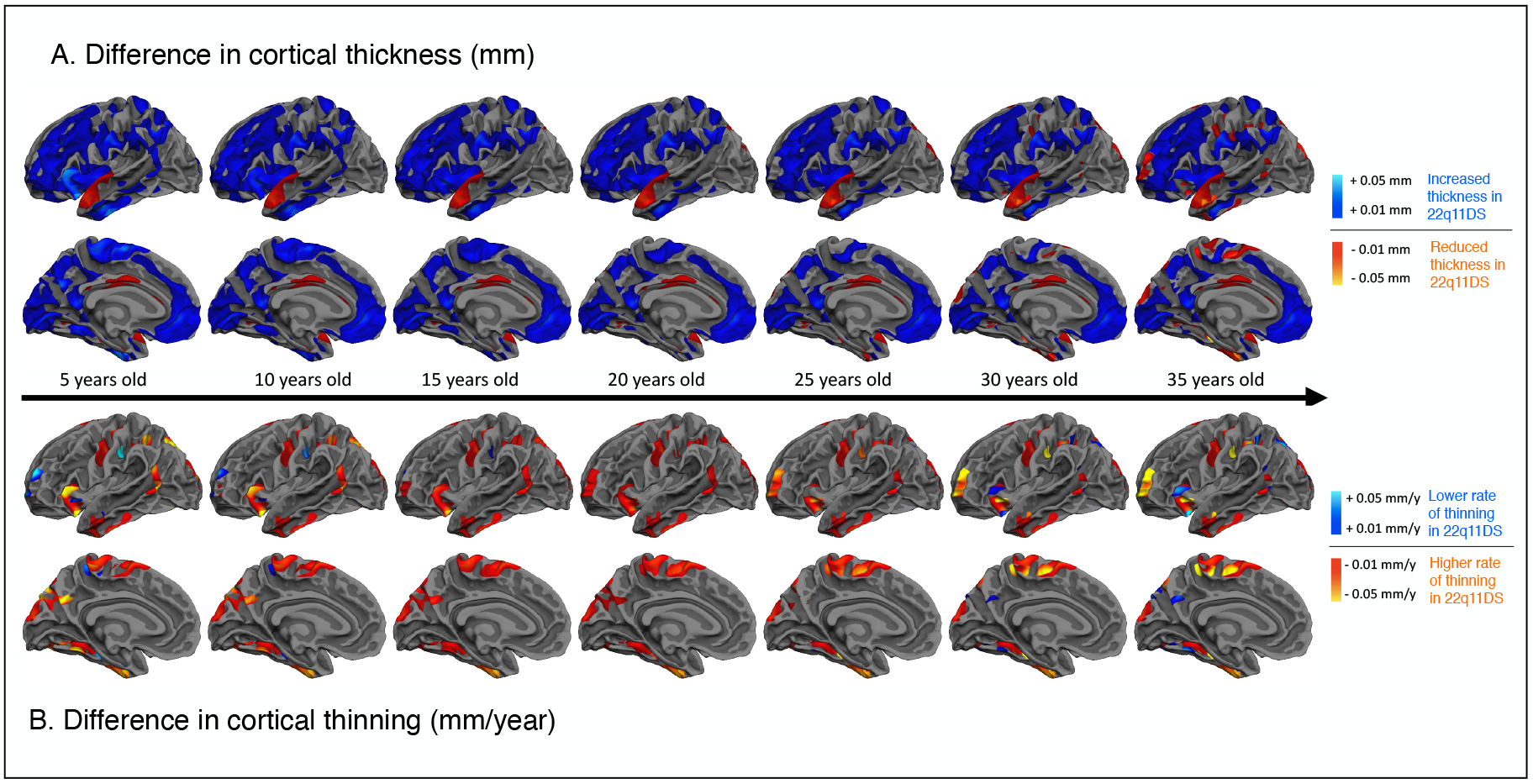
A) Differences in cortical thickness and B) differences in the annual rate of cortical thinning between 22q11DS and controls in the left hemisphere. Cold colors reflect increased cortical thickness (A) or lower thinning rates (B) in 22q11DS compared to controls; warm colors indicate reduced cortical thickness (A) or higher thinning rates (B) in 22q11DS compared to controls. While cortical thickness is increased throughout the cortex in 22q11DS, some differences tend to disappear through adolescence and into adulthood. Focal reductions in the posterior cingulate and superior temporal gyrus, however, remain present throughout development. Individuals with 22q11DS further show accelerated rates of thinning, most markedly in fronto-temporal regions. Of note, thinning rates become more pronounced in certain regions when entering adulthood (e.g., left rostral middle frontal gyrus, superior frontal gyrus, paracentral gyrus, insula, supramarginal gyrus, superior parietal gyrus, fusiform gyrus, inferior temporal gyrus), reflecting a continued thinning process in individuals with 22q11DS. Videos displaying the evolution of cortical thickness and thinning rate differences between 22q11DS and controls over time are available in the Supplementary Materials (Supplementary Video 1 for cortical thickness differences; Supplementary Video 2 for thinning differences).

### Exacerbated thinning in the STG in individuals with psychotic symptoms

Vertex-wise mixed models regression analyses comparing CT trajectories between LA-PS and N-PS participants within 22q11DS yielded a majority of linear model fits, with some constant model fits in inferior and precentral areas.

No significant group differences were found between LA-PS and N-PS groups. Significant group by age interaction effects were observed in a cluster encompassing the right STG and extending to the Heschl’s gyrus, where LA-PS individuals displayed accelerated rates of cortical thinning compared to N-PS (Figure 5).

**Figure 5.**
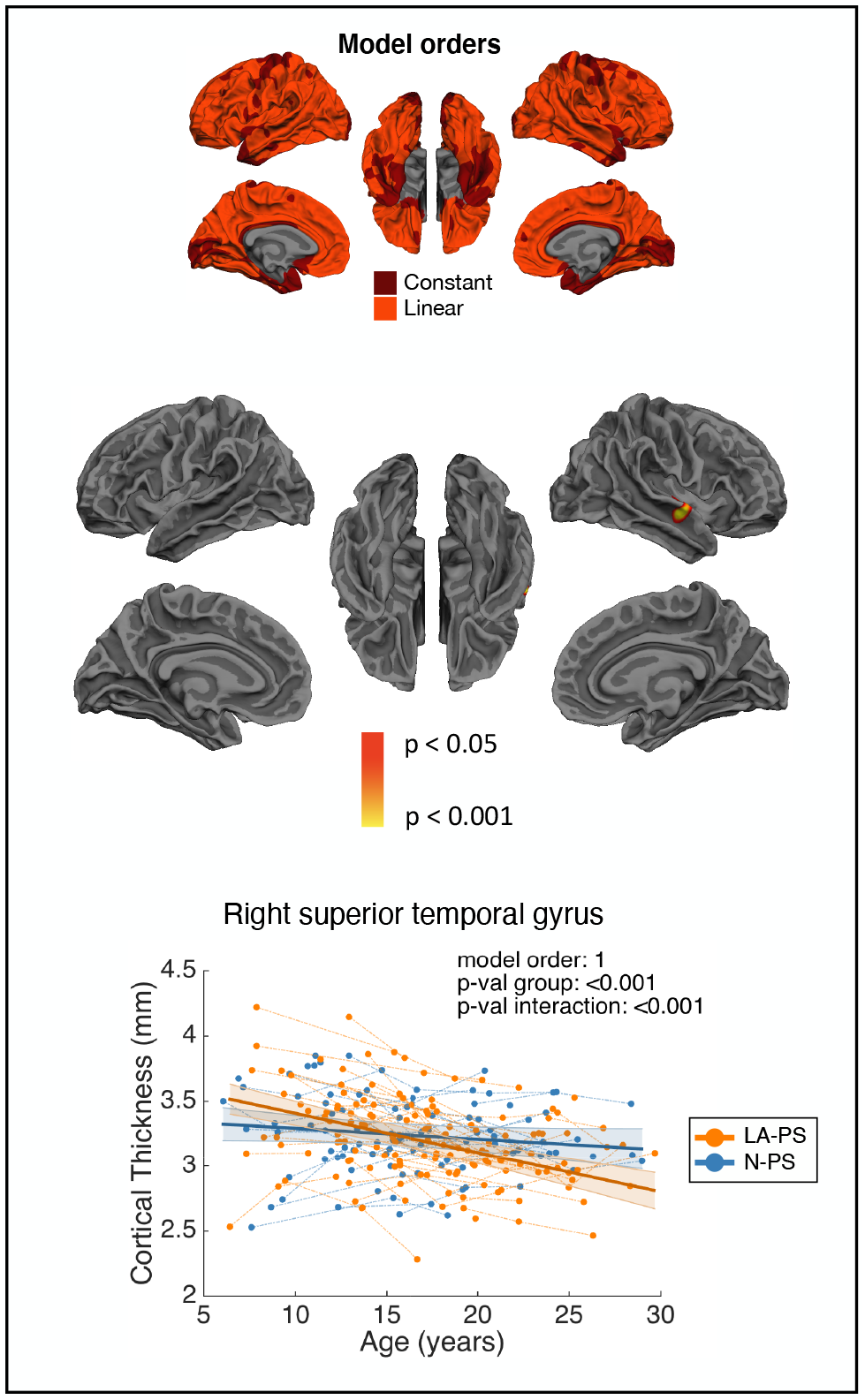
Brain maps showing regions with significant effects when comparing N-PS and LA-PS groups within 22q11DS. A significant interaction effect was found in the right superior temporal gyrus (STG), where individuals with positive psychotic symptoms showed steeper cortical thinning compared to individuals without psychotic symptoms. The upper right map represents model orders fitted at each vertex, with dark red indicating constant models and orange corresponding to linear models.

Differences in CT and in annualized cortical thinning rates between LA-PS and N-PS individuals showed mostly reduced thickness and steeper thinning rates in LA-PS participants (see Supplementary Figures S5 and S6). Supplementary videos 3 and 4 show dynamic changes over time of CT and thinning rates differences between LA-PS and N-PS groups.

To further delineate CT maturation related to the development of psychosis, we conducted an exploratory analysis comparing three subgroups within 22q11DS (N-PS, LA-PS without psychosis and LA-PS with psychosis). Interestingly, significant group by age interaction effects were again observed in right STG, where LA-PS individuals with psychosis showed even steeper thinning trajectories compared to N-PS and LA-PS without psychosis (Supplementary Figure S7). An exacerbated thinning trajectory was further found in the left lateral occipital gyrus, and CT was significantly reduced in the right superior frontal gyrus for the LA-PS with psychosis group. Supplementary Tables S3 and S4 provide detailed results for the two and three group analyses within 22q11DS.

## Discussion

This study represents the largest longitudinal investigation of CT maturation in 22q11DS to date, providing a robust delineation of CT trajectories from childhood to adulthood with an exquisite spatial and temporal resolution. Overall, findings of the current study confirm alterations generally identified in 22q11DS (19–22), and provide significant new insights into the evolution of CT over time in 22q11DS, particularly during the onset of psychotic symptoms. More specifically, vertex-wise mixed models regression analyses of CT development revealed globally decreasing CT trajectories throughout childhood and adolescence for both participants with 22q11DS and controls, with a majority of linear and quadratic trajectories. A widespread pattern of higher CT was observed in individuals with 22q11DS, with the notable exception of focal reductions in the PCC and STG. Moreover, several predominantly fronto-temporal areas displayed deviant trajectories characterized by higher CT during childhood in participants with 22q11DS compared to controls, followed by accelerated rates of cortical thinning during adolescence resulting in CT values similar to, or lower than those of adult controls. Analyses comparing N-PS *vs* LA-PS groups within 22q11DS further revealed steeper thinning rates in the right STG of participants experiencing positive psychotic symptoms. Exploratory analyses additionally suggested that thinning rates in the right STG were most exacerbated in individuals with LA-PS and overt psychosis.

In line with recent surface-based studies in 22q11DS (21–23), our longitudinal analysis confirmed the presence of widespread thicker cortex in 22q11DS. On a neurobiological level, abnormally increased CT may result from disruptions in early, prenatal developmental events involving cell proliferation and/or migration. Given that CT is thought to reflect cell density within cortical columns (37), increased CT may indicate an over-proliferation of intermediate progenitor cells during corticogenesis. Furthermore, post-mortem examinations have reported a high incidence of periventricular heterotopias suggestive of abnormal neuronal migration (38), and abnormal migration of interneurons has been confirmed in a mouse model of 22q11DS (39). 22q11DS was further characterized by focal reductions in the bilateral PCC and anterior STG, a finding that was observed in several previous studies in 22q11DS (19,21,22). Interestingly, reductions in these regions have been specifically observed in syndromic individuals with full-blown psychosis (22,40) as well as in idiopathic schizophrenia (41,42), suggesting that thinner cortex in these regions may be of specific relevance for the emergence of the illness. In a recent study on a partially overlapping sample, we showed that in individuals with 22q11DS experiencing auditory hallucinations, functional hyperconnectivity between the anterior STG and the medial geniculate nucleus (MGN) of the thalamus was associated with MGN atrophy (43). Our current findings suggest that anatomical alterations in the anterior STG may also be potentially involved. However, the exact relationship between anterior STG CT reductions and MGN atrophy in participants with auditory hallucinations remains to be determined.

Regarding trajectories of cortical development, our finding of widespread cortical thinning from childhood to adulthood is in line with recent reports on typical neurodevelopment, who describe continuous thinning throughout early childhood and adolescence (for a review, see (44)). Cortical thinning during adolescence has been proposed to reflect synaptic pruning, that is, the elimination of less used synapses and the refinement of neuronal network connections. More recently, studies have demonstrated that other mechanisms including myelination (causing the apparent grey-white matter boundary to shift outwards) and morphological changes (causing the neuropil to “stretch”), or a combination of these mechanisms, may possibly also underlie cortical thinning captured by structural MRI (24,45). For a detailed discussion on potential genetic mechanisms involved in abnormal brain development in 22q11DS, see (46).

These findings are especially interesting given the presence of accelerated rates of cortical thinning during adolescence and early adulthood in 22q11DS compared to controls found in our study. Supporting evidence for aberrant synaptic plasticity in 22q11DS comes from corresponding Lgdel mouse models of the syndrome, who have shown impaired synaptic spine stabilization (47) and reduced synaptic plasticity in parvalbumin-expressing interneurons, which have been involved in the neurobiology of schizophrenia (48). Recent longitudinal studies have also provided evidence for potentially excessive myelination (49) and aberrant development of structural morphology in 22q11DS (25). Therefore, it is possible that the genetic vulnerability for psychosis conveyed by the deletion involves anomalies in multiple tightly regulated developmental mechanisms.

Accelerated rates of cortical thinning in 22q11DS were mostly found in fronto-temporal regions, in line with previous findings in 22q11DS (23) and further confirming a growing body of evidence indicating that fronto-temporal regions are critically involved at all stages of schizophrenia, including ultra-high-risk (8,50–52), first-episode psychosis (9,52) and chronic schizophrenia (9). Interestingly, thinning in 22q11DS became even more deviant in adulthood in several frontal, medial temporal, and parietal structures, as well as in the insula. In these structures, individuals with 22q11DS continued to exhibit thinning, whereas CT tended to stabilize in controls during adulthood (25-35 years), suggesting that syndromic individuals may show early aging-related thinning (53). Given that aging has been less investigated in 22q11DS, it will be important to determine the cognitive and psychiatric outcomes associated with continued thinning during adulthood.

Of note, CT differences progressively disappeared as participants reached adulthood, but remained markedly increased in frontal areas (ages 30-35). This contrasts with typical neurodevelopment, where frontal regions, and the prefrontal cortex particularly, are known to undergo the most pronounced forms of pruning (54). While further human and animal studies will be needed to identify the neurobiological underpinnings of this finding, we can speculate that these regions may not undergo efficient synaptic elimination in the syndrome.

Within 22q11DS, participants experiencing positive psychotic symptoms showed steeper cortical thinning in the right STG, but not in other regions. While the specific involvement of the STG contrasts with previous reports indicating developmental alterations in frontal (26,28,29), cingulate (29) or parietal areas (25,29) in patients with 22q11DS experiencing psychotic symptoms, it is consistent with a growing body of evidence in 22q11DS suggesting a critical involvement of temporal areas in the emergence of psychosis (26,40,46). Reductions in the right STG have also been reported in meta-analyses of schizophrenia (6) and ultra-high-risk individuals who develop psychosis (55) and have been associated with positive psychotic symptoms in a multisite study on idiopathic schizophrenia (56), pointing to a critical involvement of this brain structure in the pathophysiology of the disease in both the general population and individuals with genetic vulnerability. Our exploratory three-group analysis further suggested that individuals with LA-PS who develop overt psychosis show the steepest thinning rates in this region, compared to LA-PS without psychosis and N-PS groups. Exacerbated thinning in the STG during adolescence is particularly relevant in the context of schizophrenia, as this region is involved in auditory processing (57), and auditory hallucinations are the most common form of hallucinations in schizophrenia. It is therefore possible that abnormal cortical maturation in the STG may play a role in the emergence of positive psychotic symptoms, including auditory hallucinations. These findings will however need confirmation in larger longitudinal samples.

Taken together, the current findings suggest that while global CT increases are likely a hallmark of 22q11DS caused by early developmental anomalies, localized accelerated thinning in the STG during adolescence may represent a valuable early biomarker related to the emergence of psychotic symptoms.

The main limitation of this study concerns the use of scanners with different field strengths (N=158 1.5T scans, N=478 3T scans), a frequent limitation in longitudinal cohorts where data collection spans through a large period of time. Nevertheless, we have previously demonstrated a high cross-scanner reliability in structural measures (35) and scanner was added as a covariate in the current analyses to control for the potential confounding effects. Next, while this study comprised the largest longitudinal analysis reported thus far, the sample available for the analysis of subgroups within 22q11DS was limited. Future longitudinal studies including larger samples of participants with psychotic symptoms, particularly individuals developing overt psychosis, are warranted to confirm our findings. Finally, data used in this study did not provide means to determine the mechanisms (i.e., aberrant pruning, myelination and/or morphological changes) responsible for aberrant CT in 22q11DS and in the emergence of psychosis. The investigation of brain maturation using animal models and human multimodal studies including, for instance, the T1/T2 ratio (44,58) allowing to measure intracortical myelination, will be important to understand the mechanisms underlying CT changes.

Overall, findings of the current study confirm the existence of neurodevelopmental alterations in a population at high genetic risk for psychosis and underline the importance of taking a developmental perspective when examining neuropathological mechanisms of this illness. For the first time, this study provided fine-grained delineations of developmental trajectories at high spatial resolution, revealing critical information regarding the timing of alterations and important hints towards the neuropathological mechanisms underlying the emergence of schizophrenia. While future studies are warranted to confirm these findings, current evidence suggests a combination of “early” pre- or perinatal alterations leading to thicker cortex in 22q11DS, combined with “late” maturational anomalies involving excessive thinning during adolescence, a mechanism that may specifically affect the right STG in the onset of positive psychotic symptoms.

## Data Availability

Publicly available resources for processing the imaging data and for conducting the statistical analyses. Code for mixed models regression is available at: https://github.com/danizoeller/myMixedModelsTrajectories
Requests for data access should be directed to the corresponding author.

## Funding

This work is supported by the National Center of Competence in Research (NCCR) Synapsy -The Synaptic Bases of Mental Diseases (SNCF, Grant number: 51AU40_125759), and by grants of the Swiss National Science Foundation to S. Eliez (#32473B_121996 and #234730_144260), M. Schaer (#163859) and M. Schneider (#PZ00P1_174206).

## Acknowledgments

The authors would like to thank all participants who contributed to this study, as well as their families and 22q11DS associations (Generation 22, Connect 22 and Relais 22) for their support. We further thank our colleagues who took part in data collection: Virginie Pouillard, Eva Micol, Johanna Maeder, Lydia Dubourg, Léa Chambaz, Clémence Feller, Justine Quiblier, Marina Goncalves, Fiona Journal and Léa Moreau. We thank Karin Bortolin for her help in data analysis. Finally, we thank the MRI operators at Center of Biomedical Imaging (CIBM) and François Lazeyras, Corrado Sandini and Farnaz Delavari for their help in the image acquisitions.

## Conflicts of interest

The authors report no financial relationships with commercial interests or conflicts of interest.

## Notes

### Competing Interest Statement

The authors have declared no competing interest.

### Author Declarations

Protocols of the study were approved by the Cantonal Research Ethics Commission of Geneva.

