## Supplementary Materials for "Altered cortical thickness development in 22q11.2 deletion syndrome and association with psychotic symptoms"

### Supplementary Method

#### *Participants*

Participants were recruited through parent associations and word of mouth for the Geneva longitudinal 22q11DS study since 2001 (1,2).

Descriptive statistics of sample demographics, age, gender and IQ differences between groups were computed using R (http://www.R-project.org/).

For the modeling of cortical thickness (CT) development in 22q11DS and controls, the sample consisted of 324 participants (N=148 22q11DS (73 male, 75 female); N = 176 controls (90 male, 86 female)) aged 5-35 years who contributed between 1-6 scans, resulting in a total of 636 scans (N=334 22q11DS (153 male, 181 female); N = 302 controls (143 male, 159 female), see Figure S1A). For individuals with multiple time points, mean time interval between scans was M=3. 571 years (SD=1.113), mean total follow-up time between the first and last scan of the same individual was M=6.713 (SD=3.728). There was no significant difference in gender (p=0.745) or age (p=0.319) between patients with 22q11DS and controls. There was a significant difference in full-scale IQ (p<0.001), which can be expected as individuals with 22q11DS have a full scale IQ of around 70-75 on average (3). Table S1 contains a summary of the above description of patients with 22q11DS and controls.

For the modeling of CT trajectories in subgroups of patients presenting lifetime attenuated positive psychotic symptoms (LA-PS) *vs* no positive psychotic symptoms (N-PS), the sample consisted of 108 patients with 22q11DS (N=47 N-PS (24 male, 23 female); N=61 LA-PS (32 male, 29 female)) aged 6-28 years who contributed between 1-5 scans, resulting in a total of 244 scans (N=98 N-PS (48 male, 50 female); N=146 LA-PS (68 male, 78 female), see Figure S1B). For individuals with multiple time points, mean time interval between scans was M=3.89 years (SD=1.314) and mean total follow-up time between the first and last scan of the same individual was M=7.15 (SD=3.305). There was no significant difference in gender (p=0.886), age (p=0.231) or full-scale IQ (p=0.22) between N-PS and LA-PS patients. Table S1 contains a summary of the above description of subgroups of patients with 22q11DS.

##### *Neurocognitive measures*

Full-scale IQ was evaluated using age-adapted versions of the Wechsler intelligence scale (i.e. the Wechsler Intelligence Scale for Children, version III or IV, or the Wechsler Adult Intelligence Scale, version III or IV) (4–7).

##### *Psychiatric assessment*

The presence of psychiatric disorders was assessed using the Diagnostic Interview for Children and Adolescents Revised (DICA-R) (8), the psychosis supplement from the Kiddie-Schedule for Affective Disorders and Schizophrenia Present and Lifetime version (K-SADS-PL) (9), and the Structured Clinical Interview for DSM-IV Axis I Disorders (SCID-I) (10) for adult patients (starting from 18 years).

##### *Image acquisition details*

T1-weighted MRI anatomical brain scans were acquired using three different scanners: a 1.5T Philips Intera scanner (158 scans), a 3T Siemens Trio scanner (300 scans) and a 3T Siemens Prisma scanner (178 scans). The 1.5T Philips Intera scanner was used from 2002-2007, the 3T Siemens Trio scanner from 2007-2014 and the 3T Siemens Prisma scanner from 2014 up to the present date. The combination of multiple different scanners is inherent to large longitudinal cohorts such as ours where data collection has been ongoing since 19 years and is also increasingly common in large-scale multi-site studies (11–13). Similar to these studies, we have addressed this issue by including scanners as a covariate in the analyses. Importantly, previous studies have shown that cortical thickness measurements are reliable, independently of scanner manufacturer or field strength (14). Moreover, we previously quantified the difference in cortical thickness estimation at each cortical point for 20 healthy participants of this sample that underwent MRI acquisitions with the two scanners on the same day, showing very high cross-scanner consistency in 90% of the cortical surface (see Method & Supplementary Figure 1 in (15)).

For the 1.5T Philips Intera scanner, the sequence was composed of 124 coronal slices with a voxel size of 0.94x0.94x1.5mm (TR=35 ms, TE=6 ms, flip angle=45°, matrix size=256x192, field of view=24 $cm^2$). On the 3T Siemens Trio and Prisma scanners, the sequence comprised 192 coronal slices with voxel size of 0.86x0.86x1.1 mm (TR=2500 ms, TE=3 ms, flip angle=8°, acquisition matrix=224×256, field of view=22 $cm^2$).

##### *Image exclusion criteria*

All scans of good quality (e.g. no excessive motion or artifacts) and without visible anomalies were included (two participants with polymicrogyria and one participant with signs of massive perinatal stroke were excluded).

##### *Image processing details*

Anatomical segmentation of T1-weighted images was performed using FreeSurfer (http://surfer.nmr.mgh.harvard.edu) version 5.1, with the goal to reconstruct accurate three-dimensional representations of the inner and outer surfaces of the cortical mantle with sub-millimeter accuracy (16,17). Automated image preprocessing included resampling into cubic voxels, intensity normalization and skull stripping. Reconstruction of the white (gray-white boundary) and pial (gray-CSF interface) cortical surfaces used deformation algorithms based on the local intensity value, geometrical and topological constraints. The obtained cortical mesh models were subsequently used for measurements of cortical thickness (CT), defined as the distance between the white (grey-white boundary) and pial (grey-CSF interface) surfaces. At the end of the reconstruction process, CT values with a submillimeter accuracy were available at 163'842 vertices by hemisphere. Segmentation accuracy was manually reviewed and corrected where necessary.

Inter-subject comparison of CT maps was then achieved through spherical registration of the surfaces to the *fsaverage* subject included in FreeSurfer, allowing reliable point-to-point comparisons of CT across all scans (18,19). Surface-based smoothing of the thickness data was done in the template's space using a FWHM of 10 mm.

To further verify the quality of CT outputs, we obtained the standardized CT distribution at each vertex, flagging scans with a CT value of more than 3 standard deviations from the norm for a particular vertex. Scans were considered outliers if they had more than 10% of vertices meeting the above-mentioned criterion. As scans selected for the study contained a maximum of 8.9% outlier vertices, all scans were considered of sufficient quality and were included in the analyses.

#### Supplementary Tables

**Table S1.** Demographic information for 22q11DS and controls and for subgroups of individuals with 22q11DS presenting high (LA-PS) or low (N-PS) positive psychotic symptoms.

| | 22q11DS | Healthy controls | p-value | 22q11DS N-PS | 22q11DS LA-PS | p-value |
|---|---|---|---|---|---|---|
| N subjects (% female) | 148 (50.68%) | 176 (58.11%) | | 47 (48.94%) | 61 (47.54%) | |
| N with 1 visit | 58 | 99 | | 19 | 15 | |
| N with 2 visits | 36 | 44 | | 12 | 19 | |
| N with 3 visits | 25 | 20 | | 10 | 16 | |
| N with 4 visits | 17 | 10 | | 5 | 10 | |
| N with 5 visits | 11 | 2 | | 1 | 1 | |
| N with 6 visits | 1 | 1 | | 0 | 0 | |
| N scans (total) | 334 | 302 | | 98 | 146 | |
| N per scanner (1.5T/3T Trio/3T Prisma) | 75/153/106 | 83/147/72 | 0.066 | 15/45/38 | 19/72/55 | 0.827 |
| Mean FSIQ | 71.24 ± 12.42 | 110.75 ± 13.32 | < 0.001 | 72.83 ± 12.12 | 71.90 ± 14.21 | 0.22 |
| Age range | 5.40-34.82 years | 5.11-32.35 years | | 6.06-28.43 years | 6.43-25.45 years | |
| Mean age | 16.47 ± 6.37 | 15.96 ± 6.56 | 0.319 | 13.54 ± 4.64 | 13.86 ± 4.29 | 0.231 |
| Mean age at first visit | 13.69 ± 6.52 | 14.59 ± 6.93 | 0.230 | 13.54 ± 4.64 | 13.86 ± 4.29 | 0.711 |
| Mean time interval between visits | 3.58 ± 1.20 | 3.55 ± 0.97 | 0.792 | 4.08 ± 1.46 | 3.77 ± 1.22 | 0.203 |
| Total follow-up time | 7.41 ± 3.77 | 5.89 ± 3.52 | 0.008 | 7.44 ± 3.25 | 6.97 ± 3.36 | 0.556 |
| N medicated | 92 (62.16%) | - | | 31 (65.96%) | 41 (67.21%) | |
| Methylphenidate | 53 (35.81%) | - | | 24 (51.06%) | 16 (26.23%) | |
| Antidepressants | 39 (26.35%) | - | | 10 (21.28%) | 25 (40.98%) | |
| Antipsychotics | 26 (17.57%) | - | | 2 (4.26%) | 18 (29.51%) | |
| Anxiolytics | 16 (10.81%) | - | | 1 (2.13%) | 9 (14.75%) | |
| Antiepileptics | 11 (7.43%) | - | | 2 (4.26%) | 6 (9.84%) | |
| More than one type of medication | 36 (24.32%) | - | | 7 (14.89%) | 21 (34.43%) | |
| N with psychiatric diagnosis | 125 (84.46%) | - | | 36 (76.60%) | 57 (93.44%) | |
| ADHD | 70 (47.30%) | - | | 22 (46.81%) | 34 (55.74%) | |
| Anxiety disorder | 96 (64.86%) | - | | 24 (51.06%) | 52 (85.25%) | |
| Mood disorder | 63 (42.57%) | - | | 16 (34.04%) | 32 (52.46%) | |
| Psychotic disorder | 21 (14.19%) | - | | 0 | 12 (19.67%) | |
| OCD | 15 (10.14%) | - | | 4 (8.51%) | 7 (11.48%) | |
| More than one diagnosis | 88 (59.46%) | - | | 26 (55.32%) | 48 (78.69%) | |

**Table S2.** Maximum values of significant clusters after cluster-wise correction for multiple comparison of mixed models comparing 22q11DS vs controls using the *mri_surfcluster* function from FreeSurfer. Max = maximum -log10(p-value) found in the cluster. Size ($mm^2$) = surface area of the cluster. Tal(X,Y,Z) = MNI coordinates of the maximum p-value. CWP = cluster-wise p-value.

| **Hemi-sphere** | **Model effect** | **Brain Region** | **Cluster Number** | **Max** | **Size ($mm^2$)** | **TalX** | **TalY** | **TalZ** | **CWP** |
|---|---|---|---|---|---|---|---|---|---|
| Left | Group effects | Postcentral | 1 | -307.653 | 24126.94 | -54.7 | -10.6 | 29.4 | 0.0001 |
| | | Lingual | 2 | -307.653 | 6382.87 | -7.7 | -77.3 | 2.2 | 0.0001 |
| | | Isthmus cingulate | 3 | -11.695 | 2054.02 | -6.8 | -53.2 | 11.2 | 0.0001 |
| | | Superior temporal | 4 | 10.954 | 1446.48 | -50.7 | 3.4 | -17 | 0.0001 |
| | | Posterior cingulate | 5 | 9.266 | 486.58 | -4.2 | -1.1 | 33.2 | 0.0325 |
| | Interaction effects | Fusiform | 1 | 5.511 | 1610.32 | -32.4 | -9.2 | -27.1 | 0.0001 |
| | | Precentral | 2 | 5.1 | 1812.82 | -43.1 | -9.8 | 36.3 | 0.0001 |
| | | Superior parietal | 3 | 4.757 | 2130.62 | -11.5 | -88.8 | 20.3 | 0.0001 |
| | | Superior frontal | 4 | 4.537 | 630.08 | -6.6 | 2.8 | 59.5 | 0.0051 |
| | | Parahippocampal | 5 | 4.364 | 708.23 | -34.7 | -29.8 | -13.4 | 0.0024 |
| | | Lateral orbitofrontal | 6 | 3.735 | 716.08 | -29.7 | 25.6 | -8.9 | 0.0021 |
| | | Superior parietal | 7 | 3.732 | 839.86 | -32.6 | -44 | 44.6 | 0.0005 |
| | | Middle temporal | 8 | 3.437 | 623.44 | -57.9 | -57.3 | 5.9 | 0.0055 |
| | | Rostral middle frontal | 9 | 2.863 | 772.79 | -22.3 | 49.8 | -1.3 | 0.0011 |
| | | Fusiform | 10 | 2.406 | 556.31 | -35.8 | -73.1 | -8.5 | 0.0129 |
| Right | Group effects | Supramarginal | 1 | -307.653 | 28740.39 | 52.9 | -24.2 | 36.5 | 0.0001 |
| | | Lingual | 2 | -307.653 | 3796.32 | 4.4 | -81.2 | 1.2 | 0.0001 |
| | | Superior temporal | 3 | 9.897 | 874.91 | 50 | -2.2 | -10.4 | 0.0001 |
| | | Middle temporal | 4 | -9.596 | 1517.17 | 50.2 | -2.9 | -26 | 0.0001 |
| | | Posterior cingulate | 5 | 7.621 | 525.08 | 3.9 | -6.1 | 33.3 | 0.0227 |
| | | Inferior temporal | 6 | -6.272 | 1076.95 | 43.1 | -57.4 | -5.1 | 0.0001 |
| | Interaction effects | Precentral | 1 | 6.239 | 2048.93 | 44.8 | -6.4 | 31.7 | 0.0001 |
| | | Lingual | 2 | 5.206 | 896.19 | 18.6 | -76.8 | -5.8 | 0.0001 |

| | | | | | | | |
|---|---|---|---|---|---|---|---|
| Inferior temporal | 3 | 4.243 | 1152.39 | 44.4 | -12.4 | -30.9 | 0.0001 |
| Insula | 4 | 4.182 | 1217.77 | 33 | -20.4 | 12.4 | 0.0001 |
| Lateral occipital | 5 | 3.732 | 1039.62 | 27.1 | -94.1 | -4.1 | 0.0001 |
| Paracentral | 6 | 3.593 | 936.08 | 6.4 | -9.9 | 57.8 | 0.0001 |

**Table S3.** Maximum values of significant clusters after cluster-wise correction for multiple comparison of mixed models comparing N-PS *vs* LA-PS individuals with 22q11DS using the *mri_surfcluster* function from FreeSurfer. Max = maximum -log10(p-value) found in the cluster. Size ($mm^2$) = surface area of the cluster. Tal(X,Y,Z) = MNI coordinates of the maximum p-value. CWP = cluster-wise p-value.

| Hemi-sphere | Model effect | Brain Region | Cluster Number | Max | Size ($mm^2$) | TalX | TalY | TalZ | CWP |
|---|---|---|---|---|---|---|---|---|---|
| Right | Interaction effects | Superior temporal | 1 | 4.288 | 552.82 | 56.7 | -9 | -5.6 | 0.0166 |

**Table S4.** Maximum values of significant clusters after cluster-wise correction for multiple comparison using the *mri_surfcluster* function from FreeSurfer for mixed models comparing three subgroups within 22q11DS: N-PS individuals, LA-PS individuals without psychosis and LA-PS individuals with psychosis. Max = maximum -log10(p-value) found in the cluster. Size ($mm^2$) = surface area of the cluster. Tal(X,Y,Z) = MNI coordinates of the maximum p-value. CWP = cluster-wise p-value.

| Hemi-sphere | Model effect | Brain Region | Cluster Number | Max | Size ($mm^2$) | TalX | TalY | TalZ | CWP |
|---|---|---|---|---|---|---|---|---|---|
| Left | Interaction effects | Lateral occipital | 1 | 3.510 | 495.20 | -26.9 | -82.0 | -8.1 | 0.02970 |
| Right | Group effects | Superior frontal | 1 | 3.277 | 605.41 | 21.8 | 0.9 | 47.6 | 0.00840 |
| | Interaction effects | Superior temporal | 1 | 4.510 | 678.06 | 48.1 | -16.2 | -2.3 | 0.00310 |

### Supplementary Figures

**Figure S1.** Scans distribution for the exploratory analysis comparing three subgroups of patients with 22q11DS: a group without psychotic symptoms (N-PS, N=47), a group of individuals with lifetime attenuated psychotic symptoms who do not develop psychosis (LA-PS no psychosis, N=49), and a group of individuals with lifetime attenuated psychotic symptoms who develop psychosis (LA-PS with psychosis, N=12). The total sample consisted of 108 patients with 22q11DS aged 6-28 years who contributed between 1-5 scans, resulting in a total of 244 scans (N=98 N-PS; N=114 LA-PS no psychosis; N = 32 LA-PS with psychosis).

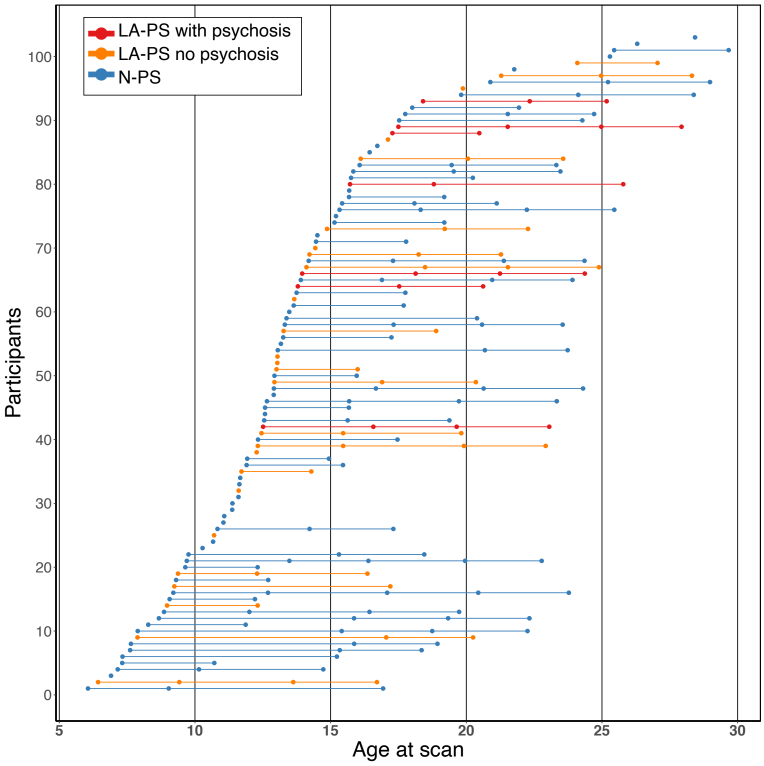

**Figure S2.** Significant intercept differences in cortical thickness between 22q11DS and controls after cluster-wise correction for multiple comparisons. In the central map, cold colors reflect increased cortical thickness and warm colors indicate reduced cortical thickness in 22q11DS compared to controls. Individuals with 22q11DS show widespread increases in cortical thickness, with focal reductions in the superior temporal gyrus and in the posterior cingulate cortex. The middle upper map depicts model orders fitted at each vertex, with dark red indicating constant models, orange corresponding to linear models and yellow indicating quadratic models.

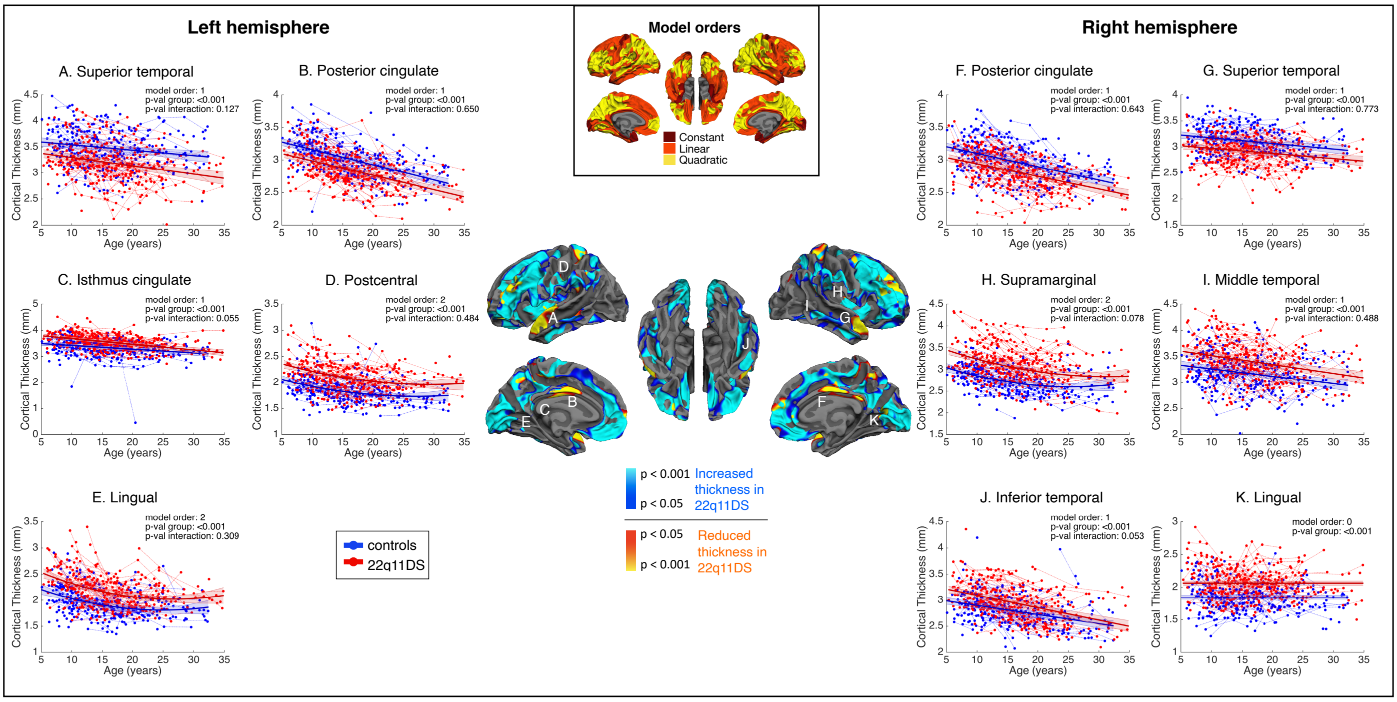

**Figure S3.** Significant shape differences in cortical thickness between 22q11DS and controls after cluster-wise correction for multiple comparisons. In the central map, warm colors reflect the degree of significance of shape differences found at each vertex. Individuals with 22q11DS show increased cortical thickness during childhood followed by accelerated thinning during adolescence, most prominently in fronto-temporal regions. The map on the lower right depicts model orders fitted at each vertex, with dark red indicating constant models, orange corresponding to linear models and yellow indicating quadratic models.

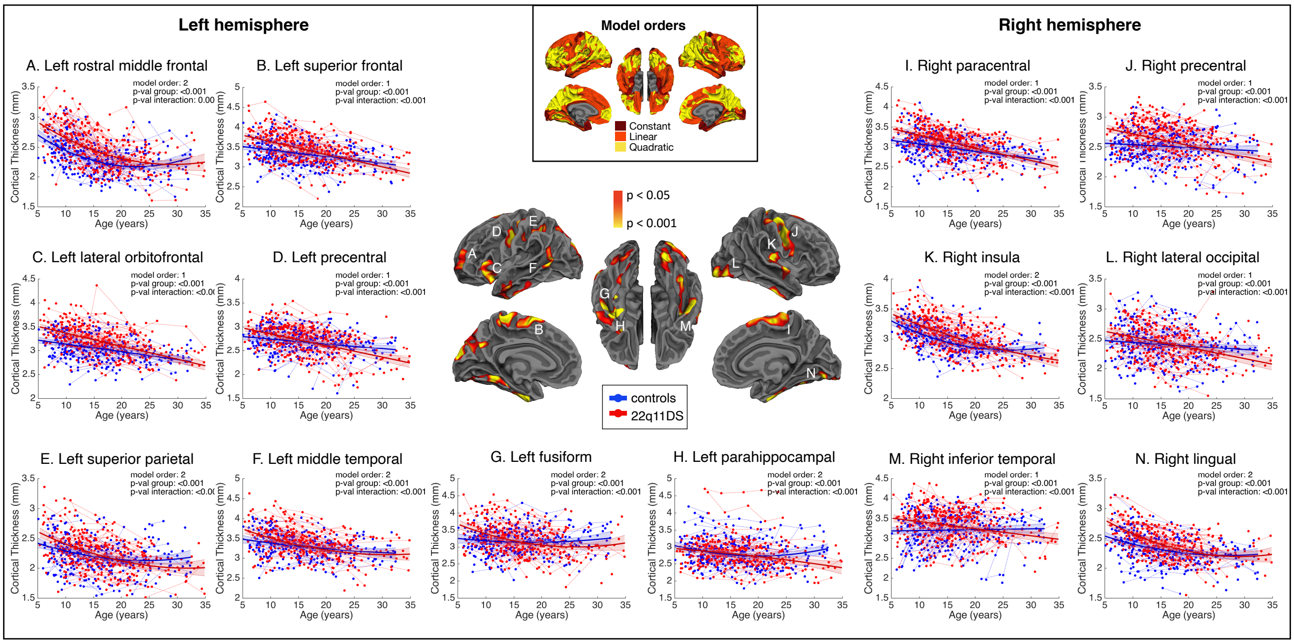

**Figure S4.** Time course displaying A) differences in cortical thickness and B) differences in the annual rate of cortical thinning between 22q11DS and controls from 5-35 years, in the right hemisphere. Cold colors reflect increased cortical thickness (A) or lower thinning rates (B) in 22q11DS compared to controls; warm colors indicate reduced cortical thickness (A) or higher thinning rates (B) in 22q11DS compared to controls. While cortical thickness is increased throughout the cortex in 22q11DS, some differences tend to disappear through adolescence and into adulthood. Focal reductions in the posterior cingulate and superior temporal gyrus, however, remain present throughout development. Individuals with 22q11DS further show accelerated rates of thinning, most markedly in fronto-temporal regions. Interestingly, thinning rates become particularly exacerbated in certain regions when entering adulthood (e.g., the right insula and lateral occipital gyrus), reflecting a continued thinning process in individuals with 22q11DS. Videos displaying the evolution of cortical thickness and thinning rate differences between 22q11DS and controls over time and in the entire brain are available in the Supplementary Materials (Supplementary Video 1 for cortical thickness differences; Supplementary Video 2 for thinning differences).

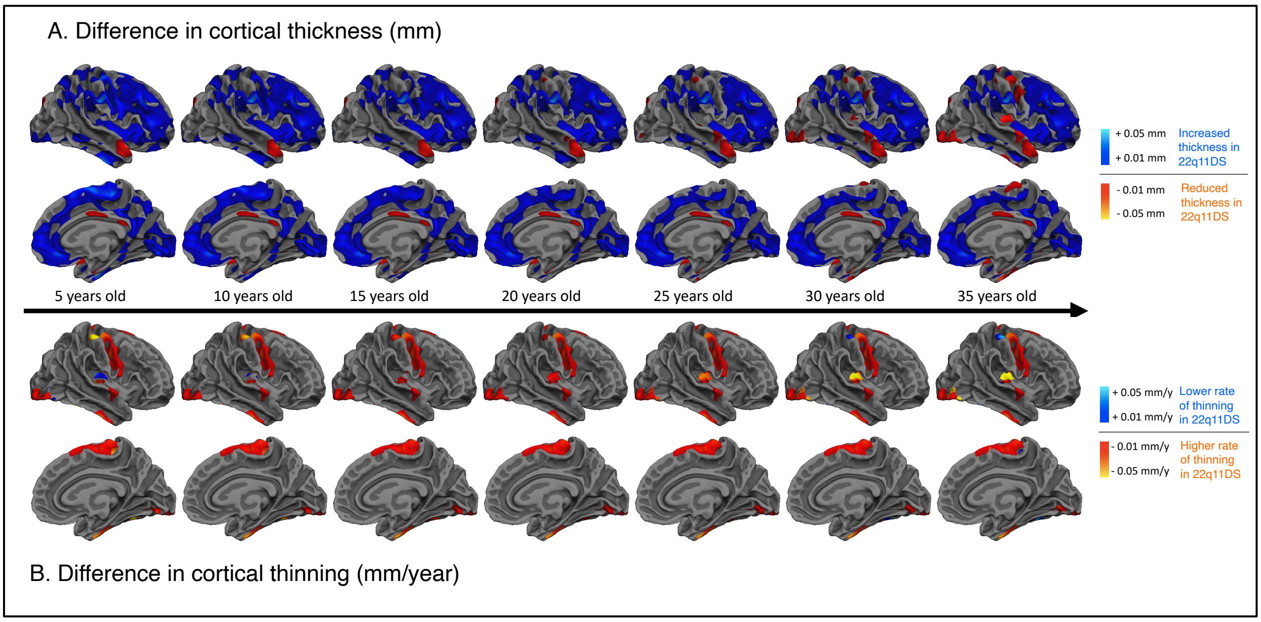

**Figure S5.** Time course displaying A) differences in cortical thickness and B) differences in the annual rate of cortical thinning between LA-PS and N-PS subgroups of individuals with 22q11DS from 6-28 years, in the left hemisphere. Cold colors reflect increased cortical thickness (A) or lower thinning rates (B) in LA-PS compared to N-PS participants; warm colors indicate reduced cortical thickness (A) or higher thinning rates (B) in LA-PS compared to N-PS individuals. Patients with positive psychotic symptoms show overall reduced CT in several frontal, temporal and parietal regions. No differences in thinning rates are observed in the left hemisphere. Videos displaying the evolution of cortical thickness and thinning rate differences between LA-PS and N-PS groups over time and in the entire brain are available in the Supplementary Materials (Supplementary Video 3 for cortical thickness differences; Supplementary Video 4 for thinning differences).

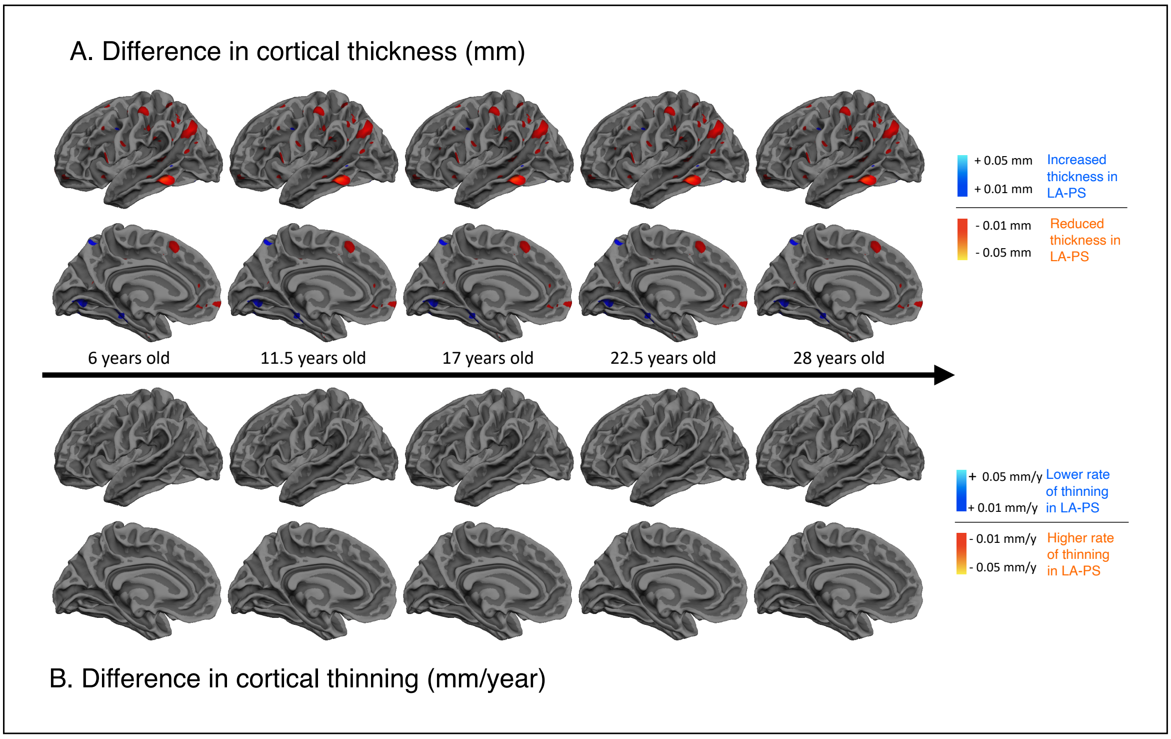

**Figure S6.** Time course displaying A) differences in cortical thickness and B) differences in the annual rate of cortical thinning between LA-PS and N-PS subgroups of individuals with 22q11DS from 6-28 years, in the right hemisphere. Cold colors reflect increased cortical thickness (A) or lower thinning rates (B) in LA-PS compared to N-PS participants; warm colors indicate reduced cortical thickness (A) or higher thinning rates (B) in LA-PS compared to N-PS participants. While individuals with positive psychotic symptoms show initially increased CT in the right STG during childhood compared to individuals without psychotic symptoms, the increases progressively disappear until showing reduced CT. Thinning rates in the right STG are steeper in individuals with psychotic symptoms and remain constant throughout development. Videos displaying the evolution of cortical thickness and thinning rate differences between LA-PS and N-PS groups over time and in the entire brain are available in the Supplementary Materials (Supplementary Video 3 for cortical thickness differences; Supplementary Video 4 for thinning differences).

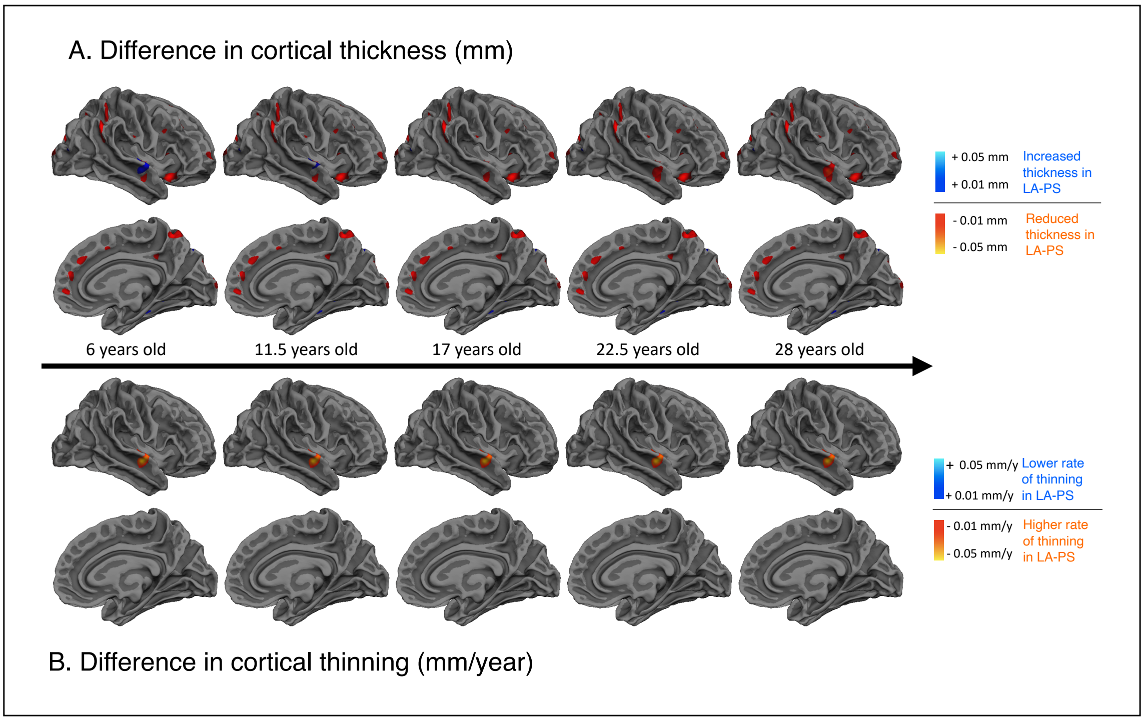

**Figure S7.** Brain maps of the exploratory analysis comparing three subgroups of individuals with 22q11DS: N-PS individuals, LA-PS individuals without overt psychosis, and LA-PS individuals with overt psychosis. In the upper left map, cold colors reflect increased cortical thickness and warm colors indicate reduced cortical thickness in LA-PS compared to N-PS patients. A significant group effect was found in the right superior frontal gyrus, where LA-PS patients with psychosis show markedly thinner cortex. In the lower left map, warm colors reflect the degree of significance of shape differences found at each vertex. Significant interaction effects were found in the left lateral occipital gyrus and right STG. In these regions, LA-PS patients with psychosis show more exacerbated thinning compared to LA-PS patients without psychosis and N-PS patients. The upper right map represents model orders fitted at each vertex, with dark red indicating constant models and orange corresponding to linear models. Results should be considered as preliminary, due to the small sample size in the group of individuals with overt psychosis.

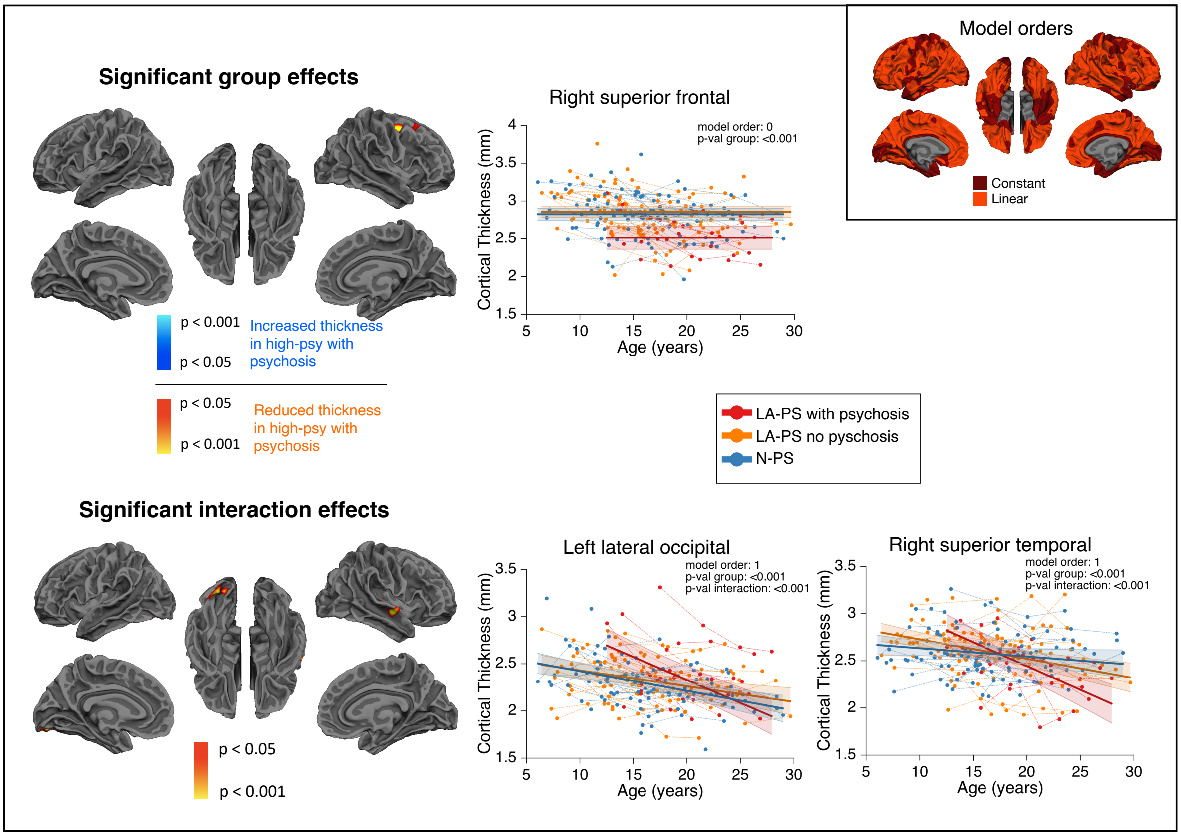
