## Supplementary Video Captions for "Altered cortical thickness development in 22q11.2 deletion syndrome and association with psychotic symptoms"

**Supplementary Video 1.** Video displaying the evolution of cortical thickness differences between 22q11DS and controls from 5 to 35 years old. Cold colors reflect increased cortical thickness; warm colors indicate reduced cortical thickness in 22q11DS compared to controls. While cortical thickness is increased throughout the cortex in 22q11DS, some differences tend to disappear through adolescence and into adulthood. Focal reductions in the posterior cingulate and superior temporal gyrus, however, remain present throughout development.

**Supplementary Video 2.** Video displaying the evolution of thinning rate differences between 22q11DS and controls from 5 to 35 years old. Cold colors reflect lower thinning rates; warm colors indicate higher thinning rates in 22q11DS compared to controls. Individuals with 22q11DS show accelerated rates of thinning, most markedly in fronto-temporal regions. Of note, thinning rates become more pronounced in certain regions when entering adulthood (e.g., left rostral middle frontal, left superior frontal gyrus, left paracentral gyrus, left supramarginal gyrus, left superior parietal gyrus, left fusiform gyrus, left inferior temporal gyrus, bilateral insula and right lateral occipital gyrus), reflecting a continued thinning process in individuals with 22q11DS.

**Supplementary Video 3.** Video displaying the evolution of cortical thickness differences between LA-PS and N-PS subgroups of individuals with 22q11DS from 6 to 28 years old. Cold colors reflect increased cortical thickness; warm colors indicate reduced cortical thickness in LA-PS compared to N-PS individuals with 22q11DS. While individuals with positive psychotic symptoms show initially increased CT in the right STG during childhood compared to patients without psychotic symptoms, the increases progressively disappear until showing reduced CT.

**Supplementary Video 4.** Video displaying the evolution of thinning rate differences between LA-PS and N-PS subgroups of individuals with 22q11DS from 6 to 28 years old. Cold colors reflect lower thinning rates; warm colors indicate higher thinning rates in LA-PS compared to N-PS individuals with 22q11DS. Thinning rates in the right STG are steeper in individuals with psychotic symptoms and remain constant throughout development.
